# Protocol for rapid implementation of a SARS-CoV-2 sero-survey during the 2020 COVID-19 pandemic – who, where, how?

**DOI:** 10.1101/2021.02.08.21251348

**Authors:** Daniel Smith, Valerie Mac, Irene Yang, Brittany Butts, Morgan Hecker, J. Christina Howell, Tugba Ozturk, Shama Pirmohammed, Hanfeng Huang, Andrea Kippels, Glenna Brewster, Danielle D Verble, Winnie Jacobs, William T. Hu, Whitney Wharton

**Author notes:** **Correspondence:** Daniel Smith.

## Abstract

**Introduction:** The 2019 novel coronavirus disease (COVID-19) pandemic has had devastating consequences in the US, yet clinical research on its natural history and transmission outside hospitalized settings has faced tangible and intangible challenges due to uncertainty in testing, case ascertainment, and appropriate safety measures. To better understand temporal evolution of COVID-19 related serological and other immune responses during a pandemic, we designed and implemented a baseline cross-sectional study of asymptomatic community volunteers and first responders in metro-Atlanta before the predicted infection peak in 2020.

**Methods:** We recruited healthy community volunteers and first responders for health history, serology, and biobanking. Through an iterative process, we identified one location on our campus and one community location which were accessible, vacant, distant from COVID-19 testing sites, open for social distancing, private for informed consent, and operational for sanitation and ventilation. Research and cleaning supplies were obtained from other researchers and private online vendors due to shortages, and faculty directly participated in consenting and phlebotomy.

**Results:** A total of 369 participants completed the study visits over six full and three half days. Over half of Phase 1 (174/299, 58.2%) and Phase 2 (45/70, 64.3%) self-reported as healthcare workers, and there was a high percentage of participants reporting exposure to known COVID-19 cases (48.2% and 61.4%).

**Conclusions:** Rigorous prospective clinical research with informed consents and is possible during a pandemic. Effective recruitment for moderately large sample size is facilitated by direct faculty involvement, connections with the community, and non-financial support from colleagues and the institution.

## 1 Introduction

The 2019 novel coronavirus (COVID-19) quickly grew into a global pandemic (1), with cases continuing to rise in the US (2). COVID-19, caused by SARS-CoV-2, can induce symptoms ranging in phenotype (e.g. cough, diarrhea, loss of smell) and severity (e.g. cough to acute respiratory failure and death (3). Accurate tracking of spread in the US has been delayed by inconsistent case definitions and shifting diagnostic algorithms. People at greatest risk for COVID-19 include the elderly (4), those with pre-existing conditions (4), healthcare workers (5), and disenfranchised populations (6, 7). Negative health outcomes due to COVID-19 in these groups are hypothesized to stem from chronic and altered inflammation (8, 9), occupational exposure (10), social determinants of health (11), or genetics. Conducting research to investigate these factors during the pandemic is challenging due to quarantine orders, fear of infection, personal protective equipment (PPE) shortage, and evolving procedures for expedited regulatory review. As exampled by retracted articles using fabricated data from high-impact journals (12, 13) and controversies surrounding treatments of unclear benefits (14, 15), desire for rigor could give way to thirst for information in the midst of a pandemic. Therefore, there remains an urgent need for thoughtful design and implementation of prospective studies during an evolving crisis.

A major obstacle in designing a prospective study related to COVID-19 is inconsistent availability of reliable testing (16). Early shortages in reverse transcription polymerase chain reaction (RT-PCR) reagents and nasopharyngeal swabs introduced numerous commercial tests – many with limited or no performance characteristics – into clinical practice through the now revised US Food and Drug Administration’s COVID-19 Emergency Use Authorization. Many current COVID-19 tests have suboptimal sensitivity (as low as 66% for RT-PCR and 67% for lateral flow rapid serology testing; 17) and single use devices do not afford the opportunity for re-testing or longitudinal, within-subject tracking using advanced technology. Most non-hospitalized Georgia residents with upper respiratory symptoms could not reliably access SARS-CoV-2 testing until early June, and disparities in access to testing along racial and socioeconomic lines further contribute to already poor outcomes in Black/African Americans and Hispanic Americans. It thus became virtually impossible to distinguish between those with mild COVID-19 infection but did not have testing, those with symptoms suggestive of COVID-19 but did not have the infection, and those who were never exposed to SARS-CoV-2. To mitigate these challenges, which jeopardize the validity of subsequent pandemic-related investigations, we rapidly recruited a large cohort of asymptomatic individuals between April and May 2020 – before the COVID-19 peak in Georgia - for cross-sectional sero-surveillance, biobanking, and future follow-up.

The aims of our serosurvey were to…. Our sero-survey of 369 people involved two phases. Phase one recruited healthy community adults (older and middle-aged without current symptoms of influenza like illness) during campus and outpatient clinic shutdown, and phase two was a directed recruitment of first responders (firefighters, emergency medical technicians [EMTs], and police officers) at the DeKalb County Fire and Rescue. The aims of this manuscript are to present the study design; recommendations for personnel, resources and study protocol; and lessons learned during a novel pandemic marked by public uncertainty, societal closure, resource limitation, and evolving best practices.

## 2 Design

### Phase 1.

We collected survey data and blood samples from 299 healthy community volunteers on April 14-17, and April 23-24, 2020 at Emory University during the statewide shelter-in-place orders. Procedures lasted approximately 40 minutes, and included informed consent, symptom checklist of previous flu-like illness, and blood draw. Participants were consented alone or with other members of their household to minimize wait times.

### Phase 2.

On May 12-13, 2020, we collected survey data and blood samples from 70 first responders at the Dekalb County Fire Rescue Headquarters. Procedures took approximately 40 minutes. Participants were recruited in groups to reduce time away from their posts. Participant were offered the option of being consented individually or with others.

### 2.1 IRB approval

This sero-survey was approved by Emory Institutional Review Board (IRB). A pre-existing inflammation-focused biobanking protocol which included healthy adults and those with aging-related diseases was expanded to include plasma testing for novel SARS-CoV-2 serology tests developed in our (WTH) lab. Due to closure of clinical testing sites for the pre-existing protocol, new physical sites were required.

### 2.2 Recruitment of Healthy Community Volunteers (Phase 1)

Initial recruitment strategies leveraged pre-existing research relationships with community members from the Atlanta area. Recruitment teams reached out to participants from author WW’s and author WTH’s cohort studies via email to describe the SARS-CoV-2 sero-survey. Unexpectedly, a number of participants challenged their social network to participate through their own electronic means (e.g., Nextdoor, listserv) reminiscent of online, crowdsourced fundraising challenges. In response to the high number of participants who contacted us as a result of this word-of-mouth campaign, we expanded our initial recruitment phase from one to two weeks, and developed a wait list in case additional resources were allocated for expanding the cohort. Given our use of a convenience sample methodology, the only exclusion criteria were those less than 18 years of age and those *currently* having symptoms of influenza like illness.

### 2.3 Recruitment of First Responders (Phase 2)

SARS-CoV-2 poses a high occupational risk to first responders (18). Based on a long-term relationship between WTH and the DeKalb County Human Services Department, the study team connected with DeKalb County Office of Public Safety and DeKalb County Fire Rescue. DeKalb County is the fourth most populous county in Georgia and has the highest population density in the metropolitan Atlanta area. Discussions with the Fire Chief and about current COVID-19-related protocols and our study design led to a joint protocol development. First responders were scheduled on May 13 and 14, 2020.

### 2.4 Environmental design

Based on evolving knowledge of SARS-CoV-2 transmission, we sought to identify locations permitting: isolation but easy access from the general public, social distancing, privacy during consent and study procedures, and thorough decontamination. Early options were eliminated due to potential confusion with COVID-19 testing sites, inadequate privacy, parking difficulties, and the need for building ventilation and custodial operations during campus shut-down. With support from the Emory University School of Medicine (SOM) leadership, the SOM building was selected for Phase I as it provided space for social distancing, remained operational with janitorial and HVAC services, had restricted access by card keys, offered visitor parking, and recently hosted a American Red Cross blood drive (Fig 1). The University Staging Department provided tables, chairs, and privacy dividers.

**Figure 1.**
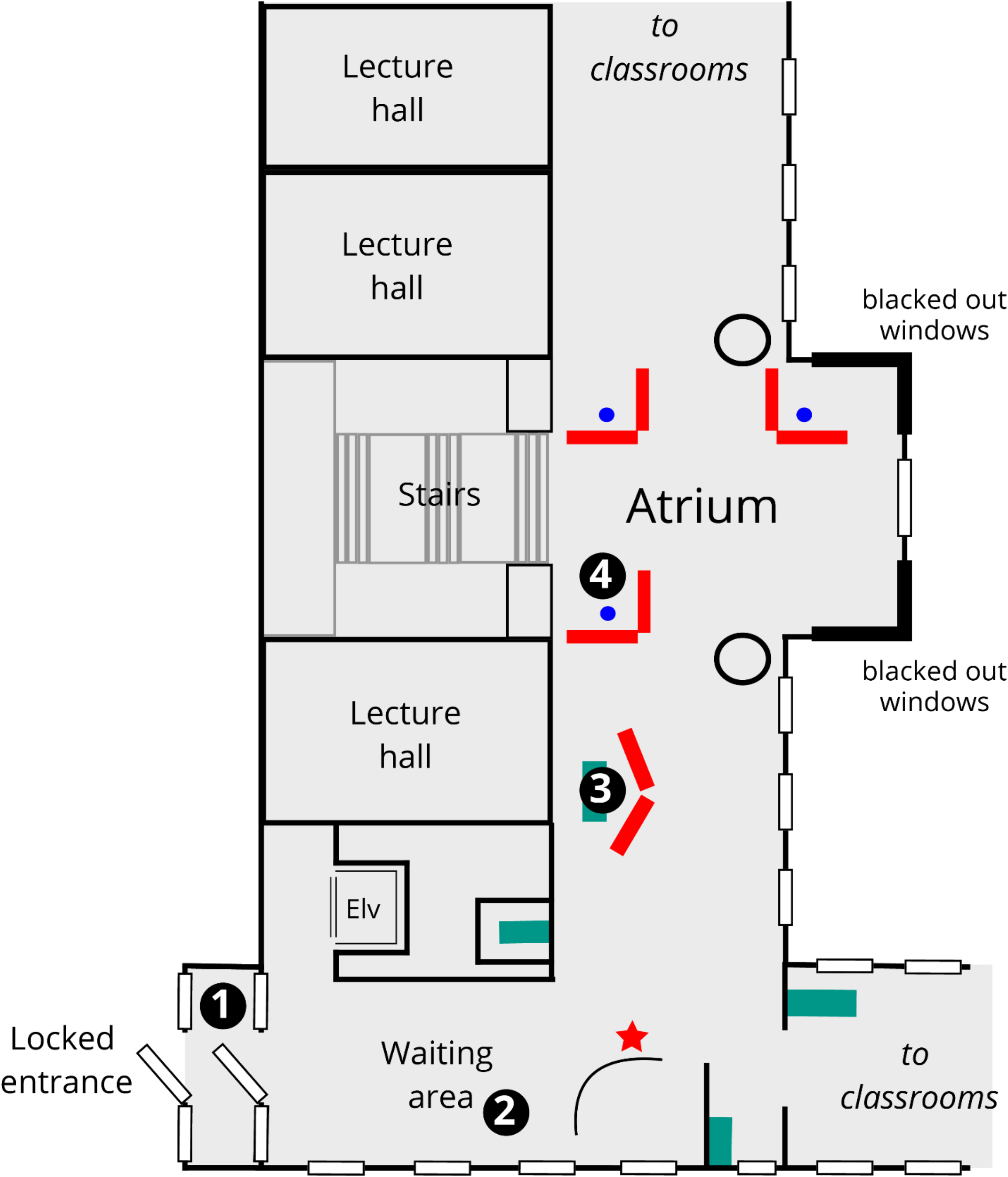
Lay-out of Emory SOM collection site. After greeter (star) saw participants outside access-controlled door, greeter proceeds to area between doors (1) to check for symptoms and take temperature using a non-contact thermometer. Participants then waited in the appropriately spaced waiting area (2) until the greeter identified an available consenter. Participants were consented individually or as a family in open but private space (3) surrounded by pre-existing walls (thick black lines) and/or privacy dividers (red). Consenters then identified available phlebotomy space before guiding participants there (4). A very small SOM staff could enter through same locked doors and access offices via elevator (elv), or through a separate entrance near the uppermost lecture hall (with a separate elevator). Windows near the phlebotomy space were blacked out for privacy.

For Phase 2, criteria similar to Phase 1 were used to at the DeKalb County Fire Rescue Headquarters. This building was closed to the public but accessible by first responders. The location was 0.7 mile from freeway ramps, and allowed for easy fire engine access to the parking lot. A large vacant classroom was converted into the study site. Participants arrived individually or as an engine crew (any crew member not participating in the study stayed in the waiting area).

### 2.5 Sero-survey Procedures

On day 1 (April 14), we piloted our study (10 participants) to assess protocol design and participant flow. Minimal changes were made following the pilot, and the first full day of data/specimens collection began on April 15.

#### Personnel

Projects were staffed by two off-site schedulers, a greeter to perform temperature and symptom check, three consenters, three phlebotomists, and additional personnel for sample transport/processing, sanitation, and stocking of supplies. Faculty (8 on-site) and staff (4 on-site, 2 off-site) from the Schools of Nursing and Medicine served on the project.

#### Equipment

Due to the scarcity of PPE at the time, participants were not provided PPE. Study team members were provided 3M N95 face masks and medical grade gloves. Faculty responsible for phlebotomy wore disposable cleanroom Tyvek suits, masks and gloves. PPE was donated by Emory School of Nursing and purchased from online vendors.

#### Standard Operation Procedures

Following best practices for field laboratory research (19), we created a standard operating procedure (SOP) for our field laboratory methods. Included in our SOPs were procedures for creating participant packets (prelabeled specimen collection tubes, two copies of the consent form, a time stamp, and a copy of the participant demographic survey). We also pre-labeled tubes for sample processing to minimize time spent in data collection areas with participants, allowing for safe and efficient sample collection. Lastly, the SOPs required specimens to be stored on ice until transported for laboratory processing within 2 hrs of collection described above.

#### Packet and Material Preparations

Preparations for sero-survey sessions required a range of preparatory tasks averaging six to eight hours per session. Study packets were created according to anticipated participant volume and included: unique research ID, consent and HIPPA forms, demographic questionnaire, COVID-19 symptom checklist, blood draw timesheet, and three, pre-labeled 10 mL K_2_-EDTA tubes. Due to high throughput (up to 80 people a day) and the need to divide samples into aliquots to avoid freeze-thawing, large numbers of microcentrifuge tubes were pre-labeled.

#### Study Visit Flow

Protocol for participant movement was governed to maintain physical distancing. Consenting and phlebotomy stations were at least 6 feet apart (Figure 1). See below for an explanation of the study visit flow:

1. A member of the study team greeted participants upon arrival and administered a verbal symptom checklist/temperature check to screen for any *current* influenza like illness before participants entered the building.
2. Upon entry, participants sat in a designated waiting area. Tables were arranged 10 feet apart. Tables and chairs were cleaned after each use with healthcare grade disinfectant.
3. Participants were then escorted to sanitized consenting stations for informed consent and collection of self-reported health histories related to past COVID-19 exposure and past symptoms of influenza like illness.
4. Participants were next escorted to a sanitized phlebotomy station.
5. Once the blood draw was completed, participants checked out with the Study PI.

#### Infection Contro

During the study visit, each consenting station (tables, chairs, and pens) was sanitized with healthcare grade sanitizing wipes in between each participant. Study personnel were expected to wear the provide 3M N95 masks at all times and were provided a new mask only if integrity of the mask was compromised. Each phlebotomy station was also sanitized with healthcare grade sanitizing wipes in between each participant. For faculty conducting phlebotomy, they were expected to put on a new pair of healthcare grade gloves with each participant. These faculty also worse disposable cleanroom Tyvek suits and, due to PPE shortages, were provided a new suit every 2 days or until the suit’s physical integrity was lost, whichever came first.

#### Laboratory Safety

Centers for Disease Control and Prevention (CDC) recommends routine diagnostic testing for specimens suspected or confirmed for SARS-CoV-2 to be conducted in a BSL-2 laboratory using standard precautions (20). However, Emory University implemented guidelines to require a BSL-2 laboratory with unidirectional airflow and BSL-3 safety precautions (PPE), also known as BSL-2+ or BSL-2 enhanced. Laboratory personnel were given SARS-CoV-2 online and in-person training, and were required to receive medical clearance.

## 3 Participant Demographics and Past Symptoms of Influenza Like Illness

### Phase 1.

299 healthy community volunteers were recruited for this phase. Mean age was 43.2 ± 12.7 years. Most participants were female (56.9%), non-Latino/Hispanic (92.5%), and White (77.3%). The vast majority of participants had ≥ 4-year college education (90.2%) and over half reported being a healthcare worker (58.2%; Table 1). Just under half reported exposure to someone with COVID-19 (48.2%) and 52% reported previously having experienced one or more symptoms associated with SARS-Cov-2. The most frequently reported symptoms were cough (74.4%), followed by nasal congestion (62.8%) and muscle aches (61.5%; Table 2).

**Table 1.** Demographic characteristics of health community volunteers (n=299) and first responders (n=70) that participated in this SARS-CoV-2 sero-survey.

|  | **Healthy Community Volunteers**<br>**(n=299)** | **First Responders**<br>**(n=70)** |
| --- | --- | --- |
|  | **n (%) or mean (SD)** | **n (%) or mean (SD)** |
| Age | 43.21 (12.69) | 41.90 (9.63) |
| Biological Sex |  |  |
| Male | 129 (43.1%) | 59 (84.3%) |
| Female | 170 (56.9%) | 11 (15.7%) |
| Ethnicity |  |  |
| Non-Latino/Hispanic | 273 (92.5%) | 67 (95.7%) |
| Latino/Hispanic | 26 (7.5%) | 3 (4.3%) |
| Race |  |  |
| African American | 33 (11%) | 15 (21.4%) |
| Asian | 30 (10.0%) | 4 (5.7%) |
| Multiracial | 3 (0.9%) | 3 (4.2%) |
| White | 231 (77.3%) | 48 (68.6%) |
| Other | 2 (0.7%) |  |
| Highest Education Completed |  | 13 (18.6%) |
| High School or less | 10 (3.3%) | 13 (18.6%) |
| 2-year College | 19 (6.4%) | 28 (40.0%) |
| 4-year College | 73 (24.7%) | 10 (14.3%) |
| Masters | 75 (25.3%) | 4 (5.7%) |
| Doctorate/Professional<br>Doctorate | 119 (40.2%) | 2 (2.9%) |
| Height (in) | 67.34 (5.44) | 69.49 (4.11) |
| Weight (lbs) | 171.16 (41.62) | 210.71 (45.80) |
| BMI | 26.24 (5.40) | 30.65 (6.33) |
| Healthcare worker |  |  |
| Yes | 174 (58.2%) | 45 (64.3%) |
| No | 125 (41.8%) | 25 (45.7%) |

**Table 2.** Self-reported exposure to SARS-CoV-2 and SARS-Cov-2 symptoms previously experienced.

|  | Healthy Community Volunteers<br>n (%) | First Responders<br>n (%) |
| --- | --- | --- |
| Known Exposure to Person with COVID-19 | 144 (48.2%) | 43 (61.4%) |
| Symptoms Experienced | N=155 | n=20 |
| Fever | 73 (47.1%) | 10 (50.0%) |
| Cough | 116 (74.4%) | 15 (75.0%) |
| Rhinorrhea/Nasal Congestion | 98 (62.8%) | 14 (70.0%) |
| Shortness of Breath | 51 (32.7%) | 9 (45.0%) |
| Muscle/Body Aches | 96 (61.5%) | 10 (50.0%) |
| Sore Throat | 81 (51.9%) | 6 (30.0%) |
| Headache | 91 (58.3%) | 9 (45.0%) |
| Vomiting | 7 (4.5%) | 3 (15.0%) |
| Diarrhea | 37 (23.7%) | 5 (25.0%) |
| Loss of smell | 22 (14.1%) | 5 (25.0%) |
| Loss of taste | 4 (2.6%) | 20 (100.0%) |
| Fatigue | 9 (5.8%) | 1 (5.0%) |
| Pneumonia | 4 (2.6%) | 19 (100.0%) |
| Chills | 2 (1.3%) | 1 (5.0%) |
| Dizziness | 3 (1.9%) | 20 (100.0%) |
| Other | 24 (15.5%) | 4 (20.0%) |

### Phase 2.

Seventy first responders were recruited for Phase 2. Mean age was 41.9 ± 9.6 years. Participants were predominantly male (84.3%), non-Latino/Hispanic (95.7%), and White (68.6%). The majority also identified as healthcare workers (64.3%; Table 1). Over half reported having had exposure to a person with COVID-19 (61.4%), and 28.6% reported previously having experienced symptoms suspected to be related to COVID-19. Notably, 100% of participants in this phase who experienced symptoms reported loss of taste, pneumonia, and dizziness. The next most frequently reported symptoms among Phase 2 participants were cough (75.0%) and nasal congestion (70.0%; Table 2).

## 4 Lessons Learned

This rapid study was accomplished through institutional space support, community partnership, and creative solutions to address logistical challenges. The following were insights gained from the implementation of our protocol:

### Institutional Space Support

One of the first lessons learned during this baseline cross-sectional study was identifying and securing a location. This study took place when the city of Atlanta, Emory University, and most out-patient Emory Healthcare operations were shut down. As such, outpatient spaces where human subject research would routinely take place were closed (except dedicated COVID-19 testing sites) or unavailable out of concerns for surface contamination. Most campus buildings did not have adequate space to facilitate social distancing or support for all-day activities. Because we recruited healthy subjects, space near campus COVID-19 testing sites– even if we utilized separate entrances – could not be secured for this sero-survey. In identifying space for human subject research across the two phases of this study, we found the ideal space must:

1. Be operational with janitorial and HVAC services
2. Have no active utilization of classrooms or closed spaces on the same floor, and have alternate access to other floors, which may be in use.
3. Accommodate participant movements to allow and encourage social distancing
4. Provide privacy for the informed consent and data collection procedures
5. Be geographically distant from active COVID-19 testing locations
6. Gain approval and support from university administration, IRB, and EHSO.

### Community Partnership

Conducting research during a pandemic highlighted the importance of strong partnerships in implementing successful community based clinical research. This was true pre-pandemic, and perhaps even more so during the COVID-19 pandemic. For example, author WW’s connections with her past participants lead to having to double our expected enrollment numbers for phase one of the study. Additionally, without author WTH’s ties with Dekalb County from non-COVID-19 projects, Phase 2 of this sero-survey would not have been possible.

### Solutions for Logistical Challenges

A major logistical hurdle of this study was the coordination of sample collection and transport to the laboratory. Whereas routine biobanking protocols allowed each study coordinator to enter the collection times into an electronic database (for calculation of time-to-freeze), such set-up was discovered to be not possible during the pilot run. Consequentially, we had to hand label specimen collection times. In order to overcome logistical challenges related to specimen collection and transport we recommend:

1. Batch-transporting samples over a 2-hr block for processing.
2. Providing copious reminders to study staff about changes in routine methodology, such as switching from electronic documentation to hand labeling specimen collection times.

We also noticed many other logistical challenges related to minimizing the time that participants were present at the study site. In order to overcome these logistical challenges, we recommend the following for sample collection and biobanking during a pandemic related shut down:

1. Creating participant packets that contain all needed paperwork for the study visit.
2. Pre-label all specimen collection tubes.

### Feasibility and Next Steps

Much discussion has taken place regarding the feasibility of randomized, double-blinded placebo-controlled clinical trials during the pandemic (28-30). A prospective observational study is typically associated with lower direct and overhead costs than a clinical trial, and outcomes from the former can bridge the gap between retrospective series and epidemiological studies to best inform natural history, disease resistance, and transmission. Our team plans to repeat the sero-survey within the same cohort in late-October/Early November 2020. This will allow us to investigate the persistence of confirmed antibody responses in this cohort, transition from early to late antibody responses, and emergence of new SARS-CoV-2 serological responses in people with no such response earlier this year.

## 5 Limitations

There were a number of unexpected developments during the execution of our study. One initial goal for Phase 1 was to recruit within communities with whom author WW had previously worked, including older adults from the metro-Atlanta LGBTQIA and Black/African American communities (21). Greater health concerns from disenfranchised and older adults during the pandemic, were not surprising to us (22, 23), but in addition to self-imposed travel restrictions, retirement communities’ mandatory quarantines limited older adults’ participation. With inconsistent information and policies across Atlanta, our recruitment goals for older adults and particularly individuals from underrepresented groups, were not met despite our team’s best effort to create a safe study site.

Another development was the result of peer-to-peer communication about our study. Word quickly spread to major Atlanta hospitals through Emory listservs and non-Emory social media sites, and a significant portion of participants subsequently self-reported as healthcare workers, subsequently increasing the education level of our sample. These workers may be more likely to commute to and from work during the shutdown than others in the city, which could increase the likelihood that they will participate in a medical research study. Given their occupational exposure (24, 25) and the strain on COVID-19 testing supplies (26, 27), it was no surprise that they wished to know their infection or serological status after working with COVID-19 patients. After our study began, we learned of two serological studies targeting healthcare workers sponsored by the US Center for Disease Control and Prevention. We thereafter referred to these other studies if we found a potential participant to work in a clinic or a hospital, but the pace of our recruitment made it challenging to verify each participant’s occupation. We do not believe deception on the participants’ part to be common or to greatly influence the outcome from our convenience sample, but it may have more significant impact on studies examining transmission dynamics and immunity durations. Future studies should prepare for these behaviors to be more common during pandemic than non-pandemic times, and accordingly adjust sample size and objective measures.

## 6 Conclusion

The COVID-19-related statewide shutdown seemingly halted clinical research overnight. However, the need for knowledge and rigor led to the creation of this sero-survey. As clinical investigators, we at times found the process to be more fragmented and reactive than pre-pandemic research. We were ultimately able to carry out a cross-sectional study of SARS-CoV-2 serology while following regulatory, ethical, and scientific principles in a modified community setting due to teamwork not only involving investigators actively guiding the study but also addressing logistical challenges by providing PPE, words of encouragement, and intellectual input. Lessons here could help others launch similar rapid prospective studies, while others – including dedication and collaboration of co-investigators who brought willingness and skillsets to the frontline – do not have easy prescriptions. Nevertheless, we show here that, with intentional and non-financial support, prospective recruitment, informed consent, and biobanking can be accomplished to add much needed scientific rigor and research ethics during a pandemic.

## Data Availability

The data generated by this study is available from the authors with reasonable request.

## 7 Author Contributions

DS: acquisition of data; analysis and interpretation of data; drafting of manuscript; critical revision of manuscript.

VM: acquisition of data; analysis and interpretation of data; drafting of manuscript; critical revision of manuscript.

IY: acquisition of data; drafting of manuscript; critical revision of manuscript. BB: acquisition of data; drafting of manuscript; critical revision of manuscript.

MH: acquisition of data; drafting of manuscript; administrative, technical or material support.

JCH: acquisition of data; drafting of manuscript; administrative, technical or material support.

TO: acquisition of data; drafting of manuscript; administrative, technical or material support.

SP: acquisition of data; drafting of manuscript; administrative, technical or material support.

HF: acquisition of data; drafting of manuscript; administrative, technical or material support.

AK: acquisition of data; drafting of manuscript; administrative, technical or material support.

GB: acquisition of data; drafting of manuscript; critical revision of manuscript.

DDB: acquisition of data; drafting of manuscript; administrative, technical or material support.

WJ: acquisition of data; drafting of manuscript; administrative, technical or material support.

WTH: conception and design; acquisition of data; analysis and interpretation of data; drafting of manuscript; critical revision of manuscript; obtaining funding; administrative, technical or material support; supervision

WW: conception and design; acquisition of data; analysis and interpretation of data; drafting of manuscript; critical revision of manuscript; obtaining funding; administrative, technical or material support; supervision

## Data Availability Statement

The data generated by this study is available from the authors with reasonable request.

